# Work-related stress and sleep problems among small-scale miners in Ghana: The role of psychological factors

**DOI:** 10.1101/2025.04.30.25326717

**Authors:** Emmanuel Nyaaba, Vanessa Francesca Epis, Lawrence Guodaar, Razak M. Gyasi

**Affiliations:** Department of Geography and Rural Development, Kwame Nkrumah University of Science and Technology, Kumasi, Ghana; African Population and Health Research Center, Nairobi, Kenya; National Centre for Naturopathic Medicine, Faculty of Health, Southern Cross University, Lismore, NSW, Australia

**Keywords:** Ghana, Mental health disorders, Sleep problems, Small-scale miners, Work-related stress

## Abstract

We examined the association between work-related stress (WRS) and sleep problems (SP) among small-scale miners in Ghana and explored the mediating roles of anxiety and depression in this association. Data were drawn from 664 miners in Obuasi. WRS and SP were assessed using the Perceived Stress Scale (PSS-4) and the WHO Disability Assessment Schedule 2.0 (WHODAS 2.0), respectively, while anxiety and depression were measured using the GAD-7 and PHQ-9. Adjusted multivariate regressions and bootstrapped serial mediation models (Hayes’ PROCESS Model 6) evaluated the hypothesized pathways. The mean (SD) age was 28.8 (8.2) years and 84.3% were males. WRS was significantly associated with SP (B = 0.2964, 95% CI = 0.2398–0.3530), with both direct (42.5%) and indirect (57.5%) effects. Anxiety mediated 38.0% of the total effect, depression 11.4%, and the anxiety–depression pathway 8.1%. Findings suggest that anxiety and depressive symptoms substantially mediate the WRS–SP relationship. Addressing occupational stress and integrating mental health support into workplace policies may enhance sleep quality and overall well-being among small-scale miners.

## 1. Introduction

Sleep quality is a key clinical and epidemiological measure linked to various health outcomes. Poor sleep has been associated with physical conditions such as obesity, metabolic syndrome, and cardiovascular disease [1,2], as well as psychological issues like cognitive decline, low mood, aggression, and depression [3–5]. Sleep problems (SP), including difficulty initiating or maintaining sleep, bad dreams, and sleep medication use, are common across occupational groups, notably among miners [6–8].

While growing evidence highlights occupational stressors as key predictors of poor sleep outcomes, one important but often underexplored factor is work-related stress (WRS). Defined by [9] as emotional, cognitive, behavioral, and physiological responses to negative aspects of work, WRS reflects a state of heightened arousal and distress. Although short-term stress responses, such as fatigue, elevated heart rate, and negative affect, are common and typically harmless, they do not disrupt sleep when the body adequately recovers during off-job periods. However, sleep may be impaired when stress recovery is insufficient and physiological systems, such as the hypothalamic-pituitary-adrenocortical and sympathetic-adrenal-medullary axes, remain activated during rest periods [10]. This prolonged activation contributes to cumulative biological strain which can ultimately result in serious health conditions [11]. WRS without adequate recovery has been shown to predict poor sleep outcomes across various populations [12,13]. For instance, a mid-life Australian cohort study found that job stress independently predicted chronic sleep disturbances [12], while a systematic review of 38 studies confirmed a consistent negative association between WRS and sleep quality [14]. Among Chinese oilfield workers, rising occupational stress correlated with increased sleep problems [15]. Despite these crucial findings from Western countries and some Asian societies, data on the association between WRS and SP in lower-middle-income countries (LMICs), particularly in the Ghanaian setting, is scarce. Crucially, while research has established clear links between occupational stress and sleep quality in formal work settings, the unique challenges faced by informal workers, especially in LMICs, remain understudied.

Importantly, the adverse effects of WRS on SP may also operate through psychological pathways. Evidence suggests that WRS contributes to increased anxiety and depression, which can disrupt emotional regulation and sleep patterns [16,17]. Anxiety, often triggered by chronic stress, can elevate arousal levels, making it difficult to initiate or maintain sleep, while depressive symptoms exacerbate these issues by influencing sleep architecture and cognitive processes related to rest [18]. Anxiety and depression frequently co-occur, with anxiety often preceding depression) [18]. While these mechanisms are well-documented in high-income countries, studies from LMICs remain limited. For instance, a study of 2,889 Chinese nurses found a significant association between occupational stress and poor sleep, mediated by anxiety and depression [19]. However, research examining the WRS–sleep problems link in LMICs, particularly among informal-sector workers such as small-scale miners, is scarce. Understanding these psychological pathways is crucial for designing targeted interventions to alleviate sleep problems and improve mental health among vulnerable workers in LMICs.

The case for examining these relationships among small-scale miners in Ghana is particularly compelling. Ghana’s small-scale mining sector employs over one million people and faces unique occupational challenges, including irregular working hours, physical demands, and exposure to hazardous conditions [20,21]. Despite ongoing policy initiatives to formalize and regulate the sector, limited attention has been paid to these workers’ occupational health and safety, particularly their mental health and sleep quality. The informal nature of their work, combined with economic pressures and regulatory uncertainties, creates a perfect storm for occupational stress. Furthermore, Ghana’s mental health infrastructure, especially in rural mining communities, remains underdeveloped, making it crucial to understand the pathways through which work stress affects health outcomes. This research gap is particularly significant given the government’s current focus on reforming the small-scale mining sector and improving occupational health standards.

In this study, the authors aim to examine the association between WRS and SP among small-scale miners in Ghana. The authors hypothesize that WRS will be linked to higher odds of SP and that this relationship will be serially mediated by anxiety and depressive (Figure 1).

**Figure 1:**
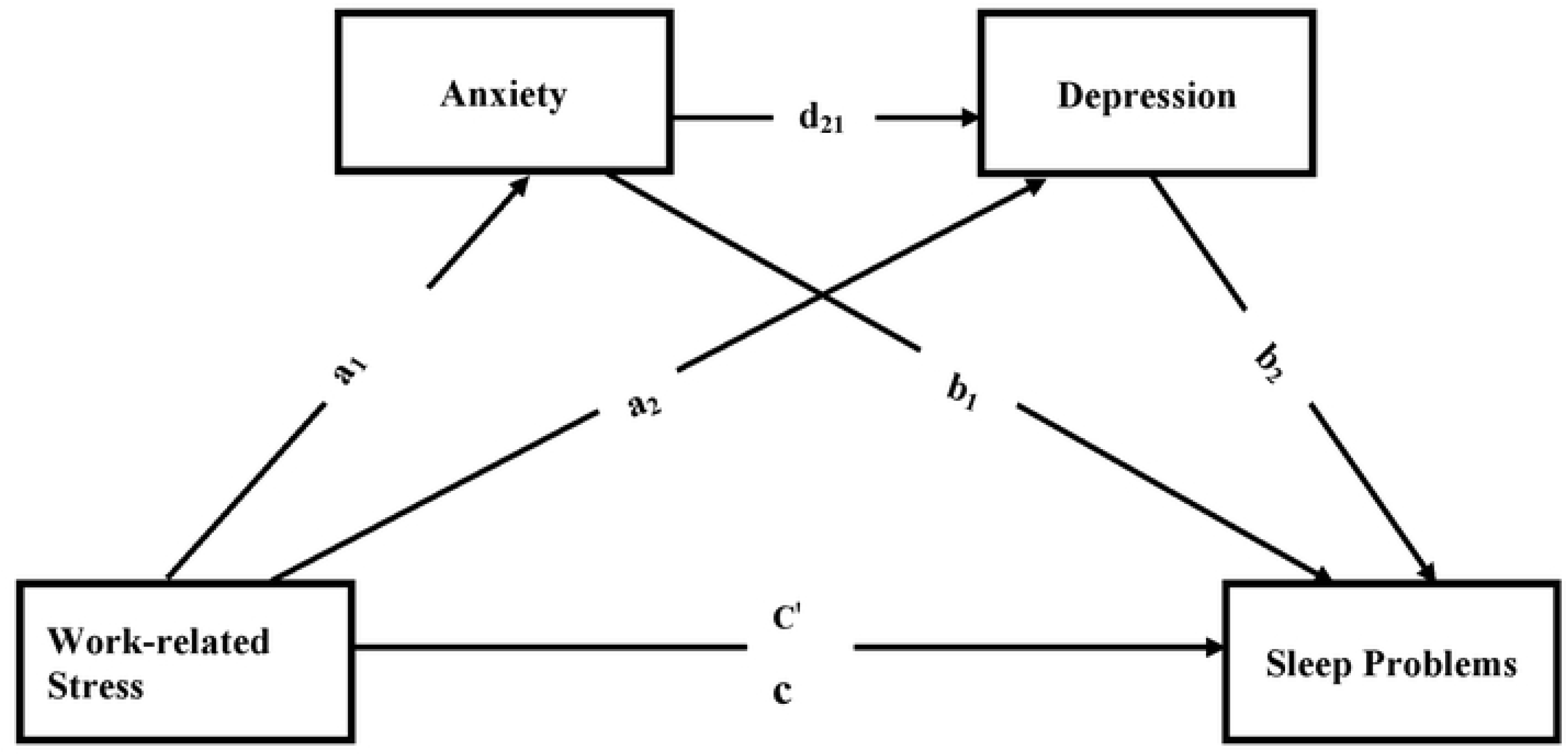
A hypothesized partial mediation model of the effect of work-related stress on sleep problems via anxiety and depression as potential mediating variables. Paths a_1_, a_2_, b_1_, b_2,_ d_21_, C’, and c, denote path coefficients. Hypothesis 1: work related stress positively affect sleep problems (total effect, c). Hypothesis 2: There is a significant specific indirect effect of work-related stress on sleep problems via anxiety (indirect effect, a_1_, b_1_). Hypothesis 3: Work related stress have a significant, specific indirect effect on sleep problems via depression (indirect effect a_2_, b_2_). Hypothesis **4:** work related stress have a significant cascading or serial mediation effect on sleep problems via anxiety and depression (serial indirect effect a_1_, d_21_, b_2_,).

## 2. Methods

### 2.1 The Survey and Participants

This cross-sectional, quantitative study was conducted in Obuasi Municipality in the Ashanti Region of Ghana, a region widely recognized for its intensive gold mining activities (Amadu et al., 2022). The Municipality comprises approximately 32 communities, most of which are noted as mining communities [22]. Out of these, seven communities—Binsere, Nhyiaeso, Gausu, Memrewa, Ntonsua, Apitisu, and Sanso, were purposively selected for this study. These communities were chosen due to their proximity to active ASM sites, the intensity of mining operations, and the prevalence of mining-related health and environmental concerns, making them ideal locations for assessing the impact of work-related stress among miners.

To ensure adequate statistical power, the study determined the required sample size using the World Health Organization’s (WHO) estimation formula [23], considering a 95% confidence interval (CI), a 5% margin of error, and a Type II error (β) of 15%, ensuring 85% power. With an assumed default prevalence (π\pi) of 0.5, the minimum required sample size was 481 participants. To account for potential nonresponse and missing data, an oversampling rate of 35% was applied, yielding a sample of 649 participants. However, the final analytic sample included 664 participants. This sample size meets the theoretical and empirical benchmarks for adequate statistical power (85%), an effect size of 0.25 [24], and robust mediation analysis [25,26]. A flow chart of the selection of the study participants is shown in Figure 2.

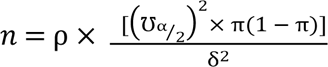

**Fig 2.**
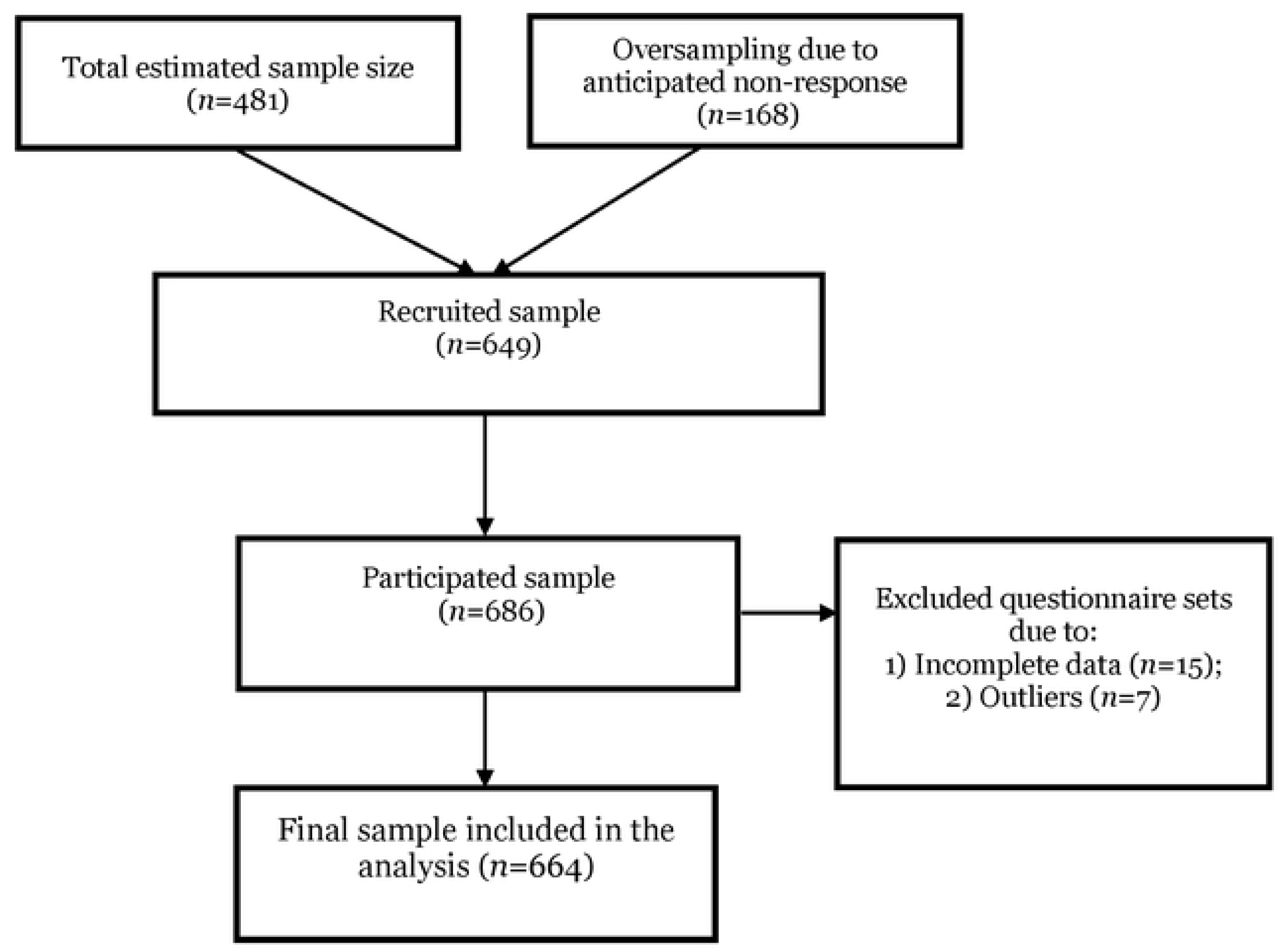
Flow chart of the selection of study participants

Participants were selected using a multi-stage sampling technique. In the first stage, the seven communities were purposively selected as described above. In the second stage, households within each selected community were sampled using a spatially guided systematic random sampling method. In smaller communities (with fewer than 250 households), every third household was selected, whereas in larger communities (250 or more households), every fifth household was chosen. In households with multiple eligible participants (i.e., miners with at least six months of mining experience), one individual was randomly selected. Additionally, artisanal miners were also recruited directly at active mining sites using a systematic interval method, selecting every fifth miner encountered during field visits. The inclusion criteria required participants to be active small-scale miners aged 18 years or older, with at least six months of continuous mining experience. Individuals who did not meet these criteria or who declined to provide informed consent were excluded from the study. To enhance efficiency and ensure timely completion, the study employed Rapid Rural Appraisal (RRA) techniques, a participatory research method designed to facilitate data collection in decentralized settings [27]. The research team was divided into four groups, each consisting of five members, ensuring a systematic and coordinated approach to reaching participants across different locations. This division allowed for effective data collection while ensuring that a larger sample size was reached within the available timeframe. This two-pronged strategy ensured that both full-time miners and those who might have been missed through household-based recruitment were included in the study. Data collection was carried out over a 8-week period (from 16^th^ February 2025 to 14^th^ April 2025)

To maintain data accuracy and reduce response bias, field teams received comprehensive training in ethical research practices, culturally appropriate engagement, and effective interviewing techniques before data collection commenced. Before data collection started, we sought ethical clearance from the Ethics Committee ***(Details and reference ID removed for double-blind review)***. All participants provided informed consent before participating in the study. Both written and verbal informed consent were obtained. Written consent was prioritized; however, verbal consent was accepted in cases where participants were unable to provide written consent due to literacy challenges. Verbal consent was documented by the research team through dated and signed consent forms, with an independent witness present to verify the consent process. No minors (under 16 years) were involved in the study; therefore, parental or guardian consent was not required. Participants were informed that their participation was voluntary, and they could choose to withdraw at any time without consequence.

### 2.2 Measures

***Work-Related Stress:*** Work-related stress was assessed using the 4-item Perceived Stress Scale (PSS-4) [28]. The PSS-4 measures perceived stress on a 5-point scale (0 = never to 4 = very often). Questions assess perceived control and ability to handle difficulties (e.g., “In the last month, how often have you felt that you were unable to control the important things in your life?”). In the current study, the variable showed acceptable internal consistency (α = 0.72).

***Sleep Problems:*** SP were measured using two items adapted from the WHO Disability Assessment Schedule 2.0 (WHODAS 2.0) [29]. Participants were asked; “Overall, in the last 30 days, 1) how much of a problem did you have with sleeping (*e.g.,* falling asleep, waking up frequently during the night, or waking up too early in the morning)? and 2) how much of a problem did you have due to not feeling rested and refreshed during the day (for example, feeling tired, not having energy)?”. Each item had five-point options from *none*=1 to *extreme*=5. Previous studies have used these items. A total score ranged from 2 to 10, with increasing scores indicating poor sleep quality (Cronbach’s alpha=.78 in this study).

***Anxiety:*** *Anxiety* was assessed using the 7-item Generalized Anxiety Disorder (GAD-7) scale [30], which has been widely used to measure anxiety symptoms in older adults. Participants were asked to recall a series of feelings from the past two weeks using seven items with four options each: 0=never, 1=for several days, 2=more than half of the days, and 3=almost every day (scores range: 0-21) with higher scores suggesting more severe anxiety symptoms. The internal consistency reliability of the scale was .79 in this study

***Depression:*** Depression was the second mediator assessed using the 9-item Patient Health Questionnaire (PHQ-9) [31]. The participants were asked to recall experiences from the past two weeks, and each item had four options: 0=rarely, 1=some days (1– 2 days), 2=occasionally (3–4 days), and 3=most of the time (5–7 days). The total score ranged 0-30; higher score reflected more depressive symptoms. The Cronbach alpha was .80 in the current study.

***Covariates***: Several demographics, health status, and occupational variables were included as control variables based on their potential influence on the study variables based on previous studies. Age was measured as a continuous variable in years. Educational level was coded on a 4-point scale (1 = no formal education, 2 = basic education, 3 = secondary education, 4 = tertiary education). Health status was assessed through chronic health conditions, including hypertension, diabetes, respiratory problems, and musculoskeletal problems, each coded dichotomously (0 = absent, 1 = present). Living arrangements were included as a proxy measure for social support, coded on a 3-point scale (1 = living alone, 2 = residing with family, 3 = living with friends/colleagues). Self-rated health (SRH) was measured using five items from the Medical Outcomes Study Short Form-36 (MOS SF-36). This included a general health rating (1 = excellent to 5 = poor), a health comparison to two years ago (1 = much better now to 5 = much worse now), and three additional items assessing perceived health status (1 = true to 5 = definitely false). A mean score was calculated, with higher scores indicating poorer self-rated health.

***Statistical analyses:*** Statistical analysis was performed using SPSS IBM version 25. Descriptive statistics were calculated for sample characteristics, with means and standard deviations for continuous variables and frequencies and percentages for categorical variables. Zero-order correlations (Pearson’s r) were conducted to examine bivariate associations between WRS, SP, anxiety, and depression. The serial mediation models were assessed using the PROCESS macro [32] with WRS as the independent variable, anxiety and depression as mediators, and SP as the outcome. Bootstrapping with 5,000 samples was used to estimate indirect effects and confidence intervals (CIs). Models were adjusted for age, sex, marital status, education, self-rated health, alcohol intake, and chronic conditions. Statistical significance was determined if the 95% CI for the indirect effect did not overlap zero, with a threshold set at p<0.05.

## 3. Results

### Sample Characteristics

The study participants (see table 1) had a mean age of 28.82 years (SD=8.17) and were predominantly male (84.3%) of the sample. The majority of the miners were single (65.1%), followed by married (24.1%), while smaller proportions were cohabiting (7.2%) or divorced/separated (3.6%). Regarding education, most miners had basic education (41.0%) or secondary education (34.9%), while 19.3% had no formal education. The sample was fairly evenly split between native residents (48.2%) and non-natives (51.8%). In terms of living arrangements, about half (50.6%) lived alone, while 39.8% lived with family, and 9.6% lived with friends or colleagues. The average work duration in small-scale mining was 5.95 years (SD=5.30). More than half of the miners (56.6%) reported alcohol intake. Regarding self-rated health, the most significant proportion rated their health as good (30.1%), followed by fair (26.5%), while excellent (15.7%), very good (14.5%), and poor (13.3%) ratings were less common.

**Table 1.**
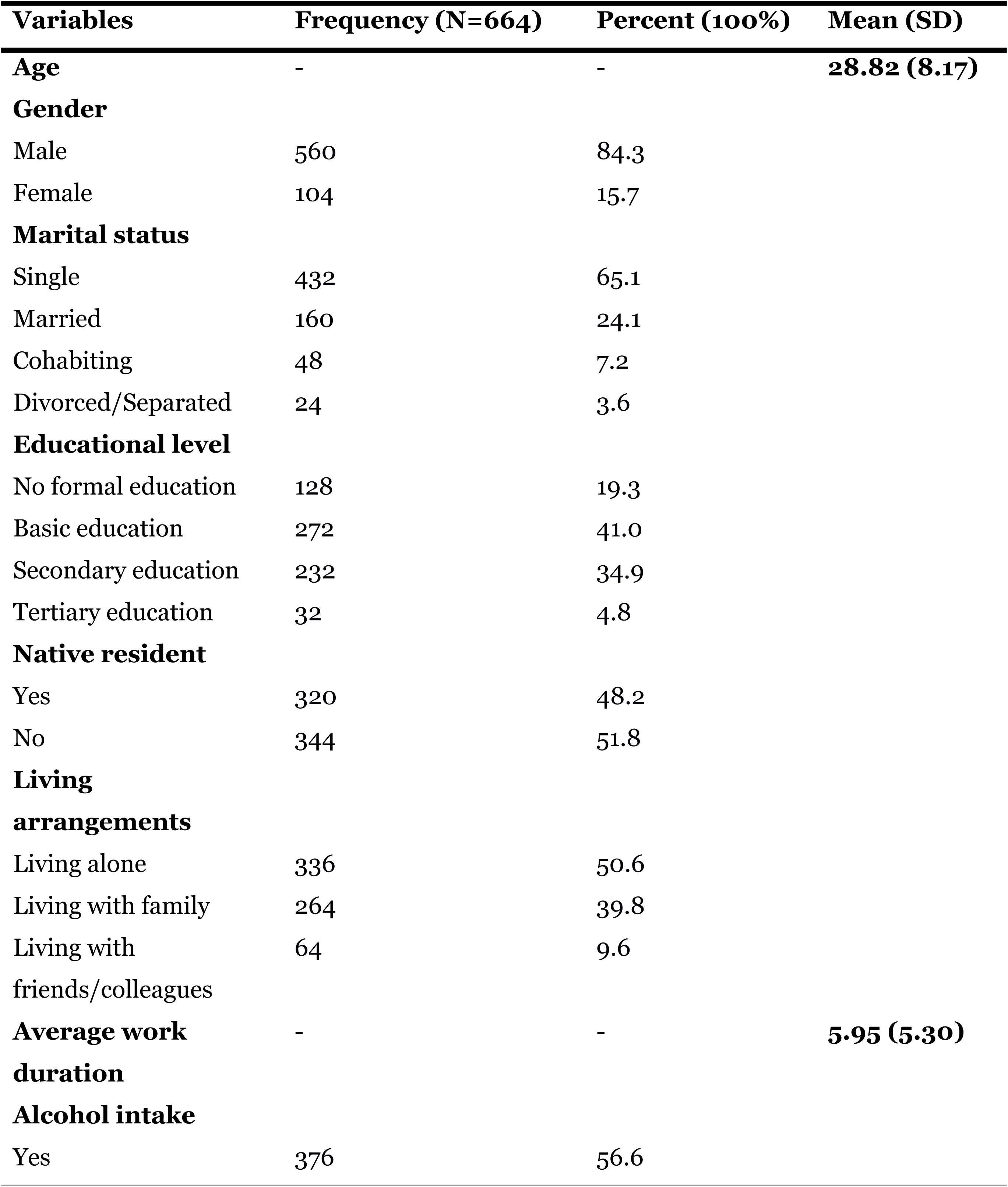
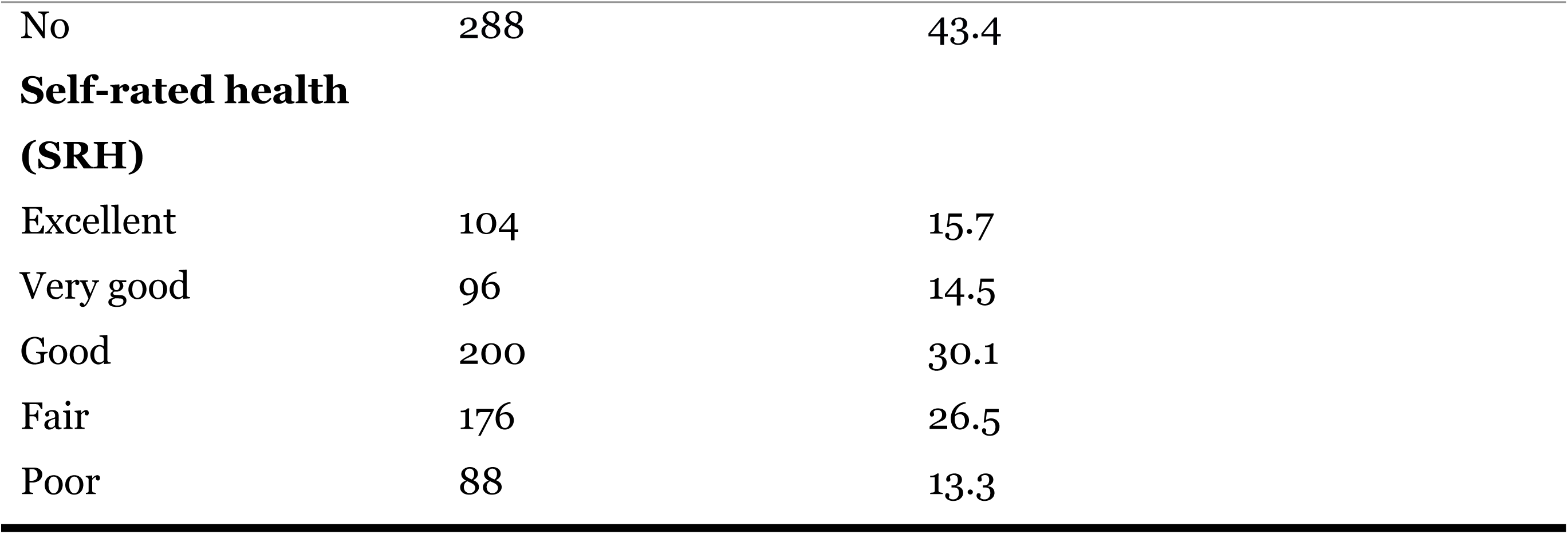
Sample Characteristics.

| **Variables** | **Frequency (N=664)** | **Percent (100%)** | **Mean (SD)** |
| --- | --- | --- | --- |
| **Age** | - | - | **28.82 (8.17)** |
| **Gender** |  |  |  |
| Male | 560 | 84.3 |  |
| Female | 104 | 15.7 |  |
| **Marital status** |  |  |  |
| Single | 432 | 65.1 |  |
| Married | 160 | 24.1 |  |
| Cohabiting | 48 | 7.2 |  |
| Divorced/Separated | 24 | 3.6 |  |
| **Educational level** |  |  |  |
| No formal education | 128 | 19.3 |  |
| Basic education | 272 | 41.0 |  |
| Secondary education | 232 | 34.9 |  |
| Tertiary education | 32 | 4.8 |  |
| **Native resident** |  |  |  |
| Yes | 320 | 48.2 |  |
| No | 344 | 51.8 |  |
| **Living arrangements** |  |  |  |
| Living alone | 336 | 50.6 |  |
| Living with family | 264 | 39.8 |  |
| Living with friends/colleagues | 64 | 9.6 |  |
| **Average work duration** | - | - | **5.95 (5.30)** |
| **Alcohol intake** |  |  |  |
| Yes | 376 | 56.6 |  |

|  |  |  |
| --- | --- | --- |
| No | 288 | 43.4 |
| **Self-rated health** |  |  |
| **(SRH)** |  |  |
| Excellent | 104 | 15.7 |
| Very good | 96 | 14.5 |
| Good | 200 | 30.1 |
| Fair | 176 | 26.5 |
| Poor | 88 | 13.3 |

### Correlations

The zero-order Pearson’s correlation matrix revealed significant positive associations between WRS and all the main study variables. Specifically, WRS was positively linked to SP (r = 0.371, p < 0.001), anxiety (r = 0.499, p < 0.001), and depression (r = 0.522, p < 0.001), as shown in Table 2. In addition, SP showed a positive correlation with both anxiety (r = 0.442, p < 0.001) and depression (r = 0.387, p < 0.001). Notably, anxiety was strongly and positively correlated with depression (r = 0.586, p < 0.001). These significant interrelations among WRS, SP, anxiety, and depression satisfied the statistical prerequisites for conducting mediation analysis.

**Table 2.** Means, standard deviations, and zero-order Pearson’s correlations between the principal study continuous variables.

| Variables | Mean (SD) | 1 | 2 | 3 | 4 |
| --- | --- | --- | --- | --- | --- |
| 1. WRS | 7.14(2.90) | 1 |  |  |  |
| 2. SP | 5.25(2.32) | 0.371*** | 1 |  |  |
| 3. Anxiety | 8.22(4.82) | 0.499*** | 0.442*** | 1 |  |
| 4. Depression | 10.36(5.69) | 0.522*** | 0.387*** | 0.586*** | 1 |

*SD—standard deviation; WRS—work-related stress; SP—sleep problems*
\*\*\* $p < .001$

### Serial mediation analysis

**Figure 3** displays the unstandardized regression coefficients for each proposed pathway. All the hypothesized models demonstrated statistical significance, even after accounting for various potential covariates through robust adjustments. The analysis revealed that WRS had a significant positive association with increased levels of anxiety (B = .8283, 95% CI = .7186–.9380), depression (B = .3620, 95% CI = .2613–.3576), and SP (B = .1259, 95% CI = .0608–.1910). Anxiety, in turn, was significantly associated with heightened depression levels (B = .3094, 95% CI = .2613–.3576) and SP (B = .1360, 95% CI = .0947–.1773). Additionally, depression was found to be significantly linked to higher SP scores (B = .0935, 95% CI = .0347–.1524). The direct relationship between WRS and SP remained statistically significant even after the inclusion of the mediating variables, suggesting that anxiety and depression serve as partial mediators in the relationship between WRS and SP.

**Figure 3.**
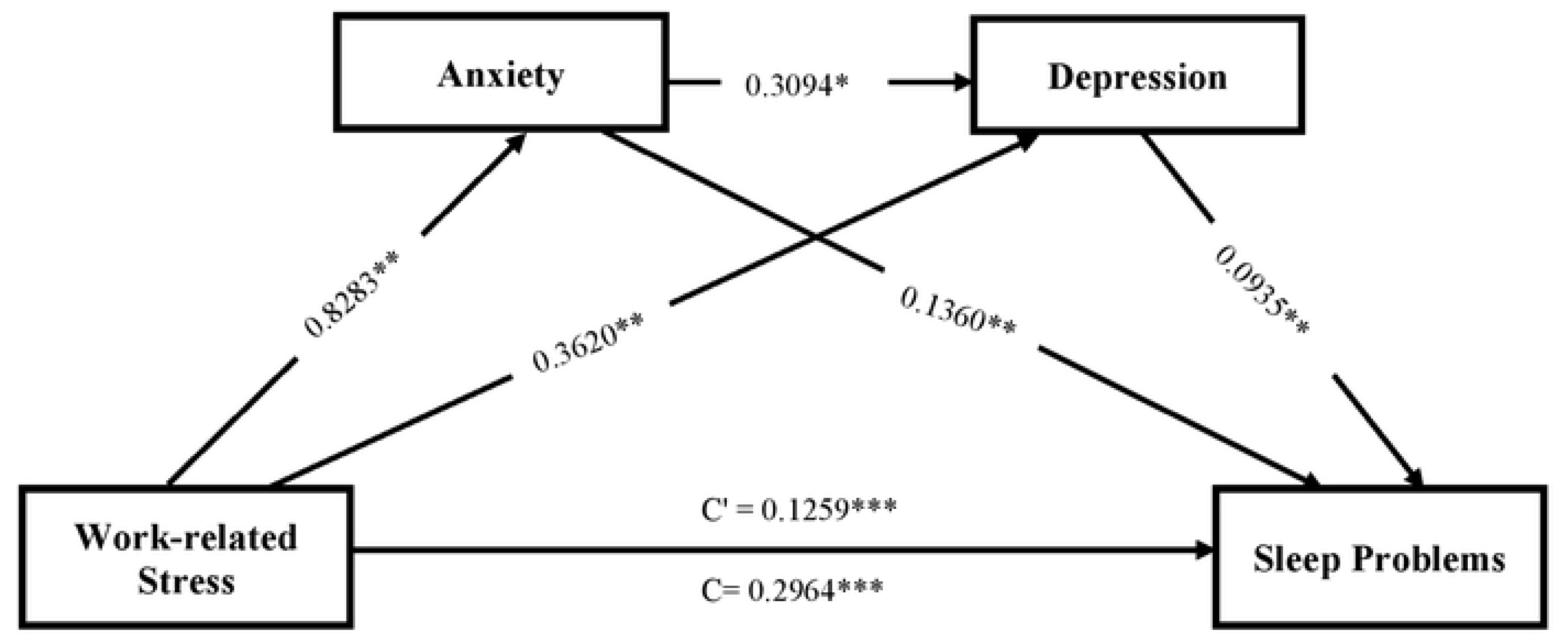
A serial multiple mediation model of the association between work-related stress and sleep problems through anxiety and depression. Path unstandardized coefficients are reported. Each model was adjusted for age, sex, marital status, educational status, living arrangement, self-rated health, alcohol intake, and chronic conditions. **p <0.005, ***p

### Bootstrapping estimates

**Table 3** presents the direct, indirect, and total effects of the relationship between WRS and SP among the gold miners in Ghana. The 95% CIs, generated using a bootstrap method with 5000 resamples, did not include zero for any of the effect estimates, indicating statistically significant indirect effects of WRS on SP through anxiety and depression, after adjusting for confounding variables. Three mediation pathways were identified: anxiety alone (B = 0.1127, BootSE = 0.0179, 95% CI = 0.0772 to 0.1471), which contributed 38.0% of the total effect; depression alone (B = 0.0339, BootSE = 0.0117, 95% CI = 0.0124 to 0.0587), accounting for 11.4%; and a serial pathway from anxiety to depression (B = 0.0240, BootSE = 0.0093, 95% CI = 0.0081 to 0.0446), which explained 8.1% of the total effect. Altogether, these indirect pathways constituted 57.5% of the overall effect of WRS on SP, while the remaining 42.5% was attributed to the direct effect.

**TABLE 3.** The indirect effects of work-related stress on sleep problems through anxiety and depression as specific and chain mediators.

| Path Model | β | BootSE | Boot 95%CI |  | % mediated |
| --- | --- | --- | --- | --- | --- |
| Direct effect | 0.1259 | 0.0332 | 0.2398 to 0.353 0 |  | 42.5% |
| *Indirect effects* |  |  |  |  |  |
| WRS -> anxiety -> SP | 0.1127 | 0. 0179 | 0.0772 | 0.1471 | 38.0% |
| WRS -> depression -> SP | 0. 0339 | 0. 0117 | 0.0124 | 0.0587 | 11.4% |
| WRS -> anxiety -> depression -> SP | 0. 0240 | 0. 0093 | 0.0081 | 0.0446 | 8.1% |
| *Total indirect effect* | 0.1705 | 0.0191 | 0.1338 | 0.2091 | 57.5% |
| *Total effect* | 0. 2964 | 0. 0288 | 0.2398 | 0.3530 | 100% |

**Note:** $\beta$ –Unstandardized regression coefficients are reported; BootsSE–Bootstrapping standard error; WRS–work-related stress; SP–sleep problems. Each model was adjusted for age, sex, marital status, educational status, living arrangement, self-rated health, alcohol intake, and chronic conditions. The empirical 95% confidence interval does not overlap with zero

## 4. Discussion

This study examined the direct and indirect pathways linking WRS to SP among small-scale miners in Ghana, using a serial mediation model. The findings revealed that WRS was independently related with higher odds of SP, with anxiety and depression explaining this link both separately and sequentially; accounting for 38.0%, 11.4%, and 8.1% of the total effect, respectively. These results underscore the need for targeted interventions that address both psychological distress and sleep health in this occupational group. Our findings align with prior research linking occupational stress to impaired sleep outcomes. For instance, Wu et al. [33] reported that among Chinese nurses, work stress affected sleep via circadian rhythm disruptions. Similarly, Yook [34] noted among 705 Korean male firefighters aged 40–50 years, occupational stress significantly correlated with cardiorespiratory fitness, arterial stiffness, and sleep quality. In the Netherlands, Van Laethem et al. [35] showed that work stress, rumination, and sleep quality were mutually reinforcing over time. Consistent with these studies, our results highlight the adverse implications of WRS for miners’ sleep health and call for integrated psychosocial interventions in the mining sector.

The present analysis identified anxiety and depressive symptoms as key psychological mechanisms mediating the relationship between WRS and SP. Anxiety accounted for approximately 38.0% of the indirect effect linking WRS to SP, suggesting that individuals experiencing heightened occupational stress are more likely to suffer sleep disturbances through increased anxiety. Consistent with previous research, chronic WRS has been associated with elevated levels of nervousness, physiological hyperarousal, and cognitive rumination, which are all hallmarks of anxiety episodes [36,37]. In hazardous work environments, such as mining and construction, continuous exposure to high demands without sufficient emotional or institutional support may trigger persistent anxiety episodes [38,39]. This heightened anxiety has been shown to impair sleep quality by increasing nighttime cortisol levels and activating the hypothalamic-pituitary-adrenal (HPA) axis [40,41]. Similarly, depressive symptoms were found to mediate approximately 11.4% of the WRS-SP association. Thus, depressive episodes serve as a secondary but meaningful pathway linking occupational stress to impaired sleep health. Prior studies have established that prolonged exposure to work-stress contributes to psychological exhaustion and low mood, increasing vulnerability to depression [35]. Depression, in turn, has been consistently linked to sleep-related complaints such as early morning awakenings, circadian rhythm disturbances, and persistent feelings of unrest [42].

This study further identified a serial mediation pathway through which WRS indirectly influenced SP via anxiety and depressive symptoms, collectively accounting for approximately 8.1% of the total effect. Consistent with existing research, our analysis showed that WRS was significantly associated with higher anxiety levels [43,44]. Over time, these anxiety symptoms appear to contribute to increased depressive symptomatology, reinforcing the mental health burden experienced by workers in high-risk and resource-constrained environments [45,46]. The co-occurrence of anxiety and depression has been found to exert a synergistic effect on sleep disturbances, with up to 80% of individuals reporting clinically significant sleep problems when both disorders are present [47]. These results suggest that the sequential accumulation of emotional distress, beginning with anxiety and progressing to depressive symptoms, undermines sleep quality through distinct yet interconnected biological and psychological mechanisms. Specifically, anxiety may initiate sleep disturbances by activating the HPA axis and heightening physiological arousal [48]. In turn, depressive symptoms may sustain and intensify these disturbances by disrupting circadian rhythms, reducing sleep drive, and impairing serotonin regulation; factors that collectively contribute to insomnia, non-restorative sleep, and poor overall sleep quality [49].

Our finding that WRS is indirectly associated with sleep disturbances through the serial mediation of anxiety and depressive symptoms (accounting for 8.1% of the variance) appears to be novel, especially within the context of small-scale miners in Ghana; a population for which this specific relationship has not been previously explored. The analysis indicates that exposure to work stress may first trigger anxiety symptoms, which subsequently heighten depressive symptoms, ultimately impairing sleep quality. This pathway aligns with prior research showing that chronic occupational stress is often linked to increased anxiety levels among workers [43,44]. Furthermore, in line with our results, earlier studies have demonstrated a strong connection between anxiety and depressive symptoms, particularly in high-pressure work environments [45,46]. The co-occurrence of these psychological conditions tends to result in more pronounced sleep issues than when either condition is present alone, with reports indicating that up to 80% of affected individuals experience substantial sleep disturbances [47]. These findings highlight the importance of addressing both anxiety and depression as intervention targets, as their combined effect on sleep involves multiple mechanisms: anxiety contributes to initial sleep disruption through hyperarousal and elevated cortisol [48], while depression induces sleep problems via altered circadian rhythms and neurotransmitter imbalances [4].

Beyond anxiety and depression tested in this study, other potential mediating mechanisms could explain the relationship between WRS and SP among small-scale miners. Musculoskeletal disorders could be an important mediator [50,51]. The physically demanding nature of mining work, combined with work-related stress, leads to musculoskeletal pain and physical exhaustion [52], operating through both physical discomfort and elevated inflammatory markers that directly interfere with sleep architecture [53]. Again, substance use behaviors, particularly alcohol consumption and stimulant use, could mediate this relationship. Research among Australian miners found that those with high occupational stress were 2.5 times more likely to engage in hazardous alcohol consumption [54], with substances significantly impairing sleep quality by disrupting sleep-wake cycles [55].

Our findings have several implications for public health, mental health care, and policy intervention in LMICs. First, healthcare providers should incorporate mental health and work-related stress screenings into routine care for miners, offering tailored interventions to address anxiety and depression that are linked to sleep problems. Second, workplace-based programs aimed at reducing stress and promoting mental well-being should be implemented. These programs could include counseling services, stress management workshops, and cognitive behavioral therapy, which are specifically designed for the mining environment. Third, awareness programs targeting miners and employers about how work-related stress leads to sleep problems through anxiety and depression can help foster a proactive approach to mental health care and reduce the stigma surrounding mental health issues. Finally, policy initiatives should focus on strengthening occupational health frameworks, including regular mental health screenings as part of health surveillance, and creating safer and more supportive work environments. Investing in organizational changes, such as reducing workplace stressors and offering support systems for miners, could help mitigate sleep disruptions and improve overall mental health outcomes.

Several strengths and limitations need to be acknowledged. First, our study is among the first to investigate the complex relationship between WRS and SP, with a specific focus on the serial mediating role of anxiety and depression among small-scale miners in Ghana; a high-risk but under-researched occupational group in the sub-Saharan African context. Importantly, we employed well-validated and widely used psychometric instruments to assess core constructs, including WRS, anxiety, depression, and SP, all with robust reliability and validity. Additionally, we controlled for a comprehensive set of potential confounders, such as demographic characteristics, health behaviors, and occupation-related variables, which enhances the internal validity of our findings. However, caution is warranted in interpreting the directionality of the observed associations due to the cross-sectional nature of the study. It remains unclear whether WRS causes SP through mental health pathways or if existing SP and mental health conditions exacerbate perceived stress levels. Future longitudinal studies are essential to clarify these temporal dynamics and disentangle the direction of causality between stress, anxiety, depression, and sleep disturbances. Also, reliance on retrospective self-report measures may introduce recall or social desirability biases. To improve robustness and generalizability, future research should consider integrating objective assessments of sleep and mental health, along with more diverse and representative samples across mining communities.

## 5. Conclusions

Using serial mediation modeling, this study provides new insights into the association between WRS and SP among small-scale miners. The findings reveal that WRS significantly impairs sleep quality, with anxiety and depression serving as key mediators. Notably, 57.5% of the WRS-SP relationship occurs through indirect pathways involving these psychological factors, both independently and sequentially. This highlights the complex interplay between occupational stress and sleep disturbances, emphasizing the need for comprehensive occupational health interventions. Effective strategies should integrate stress management programs, mental health support services, and sleep hygiene education tailored to mining workplaces. These measures could break the cycle of stress, psychological distress, and sleep problems experienced by many miners. Additionally, occupational health policies should expand beyond traditional safety concerns to prioritize mental health and sleep quality as critical components of worker well-being. We recommend future research to adopt longitudinal designs and examine diverse mining populations to confirm these findings and establish causality.

## Data Availability

All data used in this study are available from the corresponding author upon reasonable request.

## Ethics approval and consent to participate

This research adhered to the ethical principles outlined in the Declaration of Helsinki and received approval from the Committee on Human Research, Publications and Ethics (CHRPE) of the School of Medical Sciences at Kwame Nkrumah University of Science and Technology, Kumasi, Ghana (Reference: CHRPE/AP/0103/25). Informed consent was obtained from all participants prior to their participation in the study.

## Consent to participate

Informed consent was obtained from all individual participants included in the study.

## Availability of data and materials

All data used in this study are available from the corresponding author upon reasonable request.

## Authors’ contributions

**EN:** Conceptualization, Data curation, Formal analysis, Investigation, Methodology, Project administration, Visualization, Writing – original draft, Writing – review and editing. **EFV:** Data curation, Investigation, Validation, Writing – original draft, Writing, Writing – review and editing. **LG:** Supervision, Methodology, Validation, Writing – review and editing. **RMG:** Methodology, Supervision, Writing – review and editing. All authors read and approved the final manuscript

## Acknowledgements

We are grateful to all the study participants and all contributors for their support in completing this study.

